# Symptoms of depression in chronic pain: prevalence in UK Biobank and shared genetic factors

**DOI:** 10.64898/2026.04.05.26350032

**Authors:** Hannah Casey, Mark J. Adams, Andrew M. McIntosh, Marie T. Fallon, Daniel J. Smith, Rona J. Strawbridge, Heather C. Whalley

## Abstract

**Background:** Chronic pain and depression are leading causes of disability and frequently co-occur. Depression presents with diverse symptoms, but despite this variability, the prevalence of individual depressive symptoms in chronic pain and the genetic and causal associations linking these traits remain poorly characterised.

**Methods:** Using data from 142,688 age- and sex-matched UK Biobank participants, we compared depressive symptom severity levels and item-level Patient Health Questionnaire-9 (PHQ-9) prevalences, spanning affective, cognitive and somatic domains, between participants with and without chronic pain. Using genome-wide association study (GWAS) summary statistics of multisite chronic pain (MCP), major depressive disorder (MDD), and individual symptoms of depression, genetic correlations and bidirectional causal effects between MCP and depressive phenotypes (MDD and individual symptoms) were estimated via linkage disequilibrium score regression (LDSC) and two-sample Mendelian randomisation (MR), respectively.

**Results:** Depression (at every severity level) was more common in the chronic pain group compared to controls, with the largest between-group difference for severe symptoms (7.50-fold increase). All individual depressive symptoms were at least 2.79 times as prevalent in chronic pain. Additionally, chronic pain had a significant and positive genetic correlation with MDD (r_g_ = 0.59) and all depressive symptoms (r_g_ = [0.24, 0.55]). MR supported a bidirectional causal association between MCP and MDD (MCP→MDD: OR = 1.85, p_FDR_ < 0.001, MDD→MCP: β = 0.17, p_FDR_ < 0.001). At the symptom level, MR indicated bidirectional effects between MCP and anhedonia (MCP→anhedonia: OR = 1.60, p_FDR_ < 0.001, anhedonia→MCP: β = 0.08, p_FDR_ = 0.005), and unidirectional effects of MCP on appetite/weight gain (OR = 1.90, p_FDR_ = 0.022) and appetite/weight loss (OR = 1.63, p_FDR_ = 0.005), concentration problems (OR = 1.63, p_FDR_ = 0.044), and suicidal thoughts (OR = 1.46, p_FDR_ = 0.021). Additionally, genetic liability to concentration problems was associated with a lower risk of MCP (β = -0.04, p_FDR_ = 0.022).

**Conclusion:** Chronic pain is associated with a marked depressive burden spanning all symptom domains. Shared genetic architecture and symptom-specific causal pathways, particularly involving anhedonia, highlight potential targets for improved treatment of comorbid chronic pain and depression.

## Introduction

Chronic pain and depression are major global health priorities. In the Global Burden of Disease Study 2023 (Hay et al., 2025), low back pain and depressive disorders ranked first and second for years lived with disability (YLDs), across 375 diseases and injuries in 204 countries and territories. Point-prevalence estimates suggest that chronic pain and depression affect ∼20% and ∼4% of the global population, respectively (Goldberg & McGee, 2011; Organisation, 2023). Beyond their high individual prevalences, these conditions frequently co-occur, and this comorbidity is associated with greater symptom severity, poorer treatment response, and worse outcomes (Bair et al., 2003).

Previous research has demonstrated a robust phenotypic association between these conditions, with approximately 65% of depressed patients experiencing pain (Bair et al., 2003), and an estimated 50% of individuals with chronic pain meeting criteria for probable depression (Nicholl et al., 2014). Although the reasons for this frequent co-occurrence are not fully understood, a growing literature implicates shared biological mechanisms as part of the explanation (Sheng et al., 2017). It has also been proposed that the relationship is bidirectional, such that each condition can precipitate and exacerbate the other (Johnston et al., 2019; Tang et al., 2022; Zhao et al., 2023). Genome-wide association studies (GWAS) of chronic pain and depression have demonstrated significant genetic contributions in the development of both disorders (Adams et al., 2025; Johnston et al., 2019). In particular, a GWAS of multisite chronic pain (MCP) reported a substantial cross-trait genetic correlation with major depressive disorder (MDD; r_g_ = 0.53) and Mendelian randomisation (MR) analyses indicated evidence consistent with a direct effect of genetic liability to MCP on risk of MDD (Johnston et al., 2019).

Given the complexity of chronic pain, several studies have also used granular phenotyping, disaggregating by pain condition and affected body site, to demonstrate strong phenotypic (Aaron et al., 2025), genetic (Meng et al., 2020), and putative causal associations across specific pain subtypes with depression (Jiang et al., 2025; Tang et al., 2022; Wang et al., 2024). Depression, like chronic pain, is highly heterogeneous in its presentation and underlying pathophysiology. Diagnoses typically require the presence of at least one core symptom, depressed mood or anhedonia, alongside several additional symptoms that span affective, cognitive and somatic domains (American Psychiatric Association, 2013). Consequently, depression is clinically diagnosed based on multiple symptom combinations. For example, Diagnostic and Statistical Manual of Mental Disorders-5 (DSM-5) criteria permit 227 distinct symptom combinations for a major depression diagnosis (Zimmerman et al., 2015). Furthermore, recent genetic studies have shown that genetic architecture varies between individual depressive symptoms (Adams et al., 2024; Thorp et al., 2020).

Previous studies have investigated the association between individual depressive symptoms and chronic pain using observational designs (Knaster et al., 2016; McWilliams et al., 2017; Yang et al., 2022). Here, specific symptoms have been highlighted as prominent or central in chronic pain, including fatigue (McWilliams et al., 2017; Yang et al., 2022), sleep difficulties (Yang et al., 2022), anhedonia and concentration problems (McWilliams et al., 2017). While these findings offer important insights into depression symptoms in chronic pain, conclusions are constrained by modest sample sizes of these studies (typically hundreds of participants) and have yet to be extended to well-powered, symptom-level genetic analyses, leaving the genetic associations and putative causal pathways uncertain. Robustly characterising which symptoms are most strongly associated with chronic pain on an observational, genetic and causal level may not only help us understand the cause of this comorbidity but also identify targets for screening and treatment of co-occurring chronic pain and depression

Therefore, in the present study, the prevalence of individual depressive symptoms in chronic pain was estimated using UK Biobank (UKB; n = 142,688). In parallel genetic analyses, cross-trait genetic correlations and estimates of bidirectional causal associations between chronic pain and depression, as well as between chronic pain and individual depressive symptoms, were estimated by leveraging independent publicly available GWAS summary statistics for MCP, MDD and individual depressive symptoms.

## Methods

### UK Biobank sample

UKB is a large-scale, prospective cohort study that recruited approximately 500,000 participants aged 40–69 years across the UK between 2006 and 2010 (Bycroft et al., 2018). To date, the study continues to collect extensive health-related data, including demographics, lifestyle factors, physical measurements, biological samples, and genetic information. Participants regularly completed follow-up questionnaires covering a wide range of health-related outcomes, including mental health and pain experiences. To estimate the prevalence of depressive symptoms in chronic pain, we analysed complete, age- and sex-matched data from 142,688 participants who completed the online Experience of Pain (EoP) questionnaire (UKB category 154).

### Chronic pain status

In the EoP questionnaire, participants were asked whether they had been “troubled by pain or discomfort, either all the time or on and off, that has been present for more than 3 months”. Those responding “Yes” were classified as chronic pain cases, and those responding “No” were classified as chronic pain-free controls for our observational analyses. Full source details for each UKB phenotype are provided in Supplementary Table 1.

### Depressive symptom phenotypes

Depressive symptoms were assessed using the Patient Health Questionnaire-9 (PHQ-9), included in the EoP questionnaire. The PHQ-9 is a widely used, self-report instrument that assesses the frequency of nine depressive symptoms over the preceding two weeks (Kroenke et al., 2001). Each symptom is rated on a four-point Likert scale: 0 = “Not at all”, 1 = “Several days”, 2 = “More than half the days”, 3 = “Nearly every day”. A total PHQ-9 score ranging from 0 to 27 is calculated by summing responses across all nine items. In our observational analysis, depressive symptom severity was defined over four levels: mild, moderate, moderately severe and severe, corresponding to PHQ-9 score cutoffs of 5, 10, 15, and 20, respectively. For symptom-level analyses, participants were classified as cases for a given depressive symptom if they responded “More than half the days” or “Nearly every day” and those who responded “Not at all” or “Several days” were classified as controls. Details of the source of each PHQ-9 item are available in Supplementary Table 1.

### Phenotypic association analysis

All analyses were conducted in R (v4.4.1). To balance age and sex across chronic pain groups, we performed exact 1:1 age- and sex-matching using MatchIt (v4.7.2). We compared the prevalence of depressive-symptom severity tiers and individual symptoms between participants with and without chronic pain using Pearson’s χ² test (chisq.test()). We controlled the false discovery rate (FDR) at 5% using the Benjamini-Hochberg procedure across 13 tests (p_FDR_ < 0.05).

### Genetic correlation analysis

To estimate the genetic correlation between chronic pain and depression and depressive symptoms, we performed linkage disequilibrium score regression (LDSC) using LDSC (v1.0.1) (Bulik-Sullivan et al., 2015). Analyses were conducted using precomputed European LD scores from the Broad Institute (https://data.broadinstitute.org/alkesgroup/LDSCORE/eur_w_ld_chr.tar.bz2), restricted to genetic variants included in HapMap3 (https://data.broadinstitute.org/alkesgroup/LDSCORE/w_hm3.snplist.bz2). Multiple testing was controlled at a 5% FDR using the Benjamini-Hochberg procedure across 11 comparisons (p_FDR_ < 0.05). Where possible, we selected the largest available GWAS datasets in European populations that closely matched the phenotypes used in our phenotypic association analysis. To minimise bias from sample overlap, we prioritised phenotype pairs derived from independent samples wherever possible.

Major Depressive Disorder (MDD): We used summary statistics from the Psychiatric Genomics Consortium (PGC) trans-ancestry genome-wide meta-analysis of MDD comprising 357,636 MDD cases and 1,281,936 controls (Adams et al., 2025). For all analyses, we used the European-ancestry subset excluding UKB and 23andMe.

Depression symptoms: GWAS summary statistics for individual DSM-aligned symptoms meta-analysed across community cohorts (ALSPAC, Estonian Biobank, UKB-Mental Health Questionnaire (MHQ)) were used (Adams et al., 2024). Across symptoms, community cohort sample sizes ranged from 40,180 to 307,280. Valid summary statistics of depressive symptoms comparable to those present in the PHQ-9 were fully available, with the exception of psychomotor changes, as psychomotor agitation and slowing were not found to have h^2^ greater than 0 and were therefore not included in genetic analysis.

Multisite Chronic Pain (MCP): We used UKB summary statistics for MCP (count of body sites with pain > three months) in 387,649 participants (Johnston et al., 2019). To ensure independence between our genetic measures of MCP and depressive symptoms, we additionally conducted an in-house GWAS of MCP in 280,184 UKB participants who had not completed the Mental Health Questionnaire (MHQ), replicating the materials and methodology outlined by Johnston et al. Full details on each publicly available GWAS is provided in Supplementary Table 3.

### Mendelian randomisation analysis

To estimate the potential causal relationship between chronic pain and depression and depressive symptoms, we conducted a bidirectional two-sample univariable MR analysis. All MR analyses were performed in R (v4.4.1) using the TwoSampleMR (v0.6.17) and MR-PRESSO (v1.0) packages. When MCP and MDD were assessed as exposures, genetic variants meeting genome-wide significance (p < 5 × 10⁻⁸) were selected as instrumental variables (IVs). For analyses in which individual depressive symptoms were the exposure, we used a suggestive threshold (p < 5 × 10⁻^6^) to obtain a sufficient number of IVs, given the smaller number of genome-wide hits available for these traits. The use of a more liberal instrument-selection threshold is acceptable when paired with measures such as instrument-strength filtering, robust MR methods and comprehensive sensitivity analyses, as implemented in this study (Burgess et al., 2019). To ensure independence, IVs were clumped using a linkage disequilibrium (LD) threshold of r² < 0.001 within a 1000 kb window, with LD estimated from the 1000 Genomes Project Phase 3 European-ancestry reference panel (Auton et al., 2015). Exposure and outcome summary statistics were harmonised, and palindromic IVs were aligned where possible using minor allele frequency data. We reported odds ratios (OR) with 95% confidence intervals when the outcome was binary (MDD and depressive symptom phenotypes), and β coefficients with 95% confidence intervals when the outcome was continuous (MCP).

We applied multiple MR methods, including inverse-variance weighted (IVW) (multiplicative random effects), MR Egger and weighted median, each with different assumptions regarding horizontal pleiotropy. IVW served as the primary analysis method, and consistency in magnitude and direction of effect across secondary methods was used to support causal inference. However, statistical significance in these secondary methods was not required to validate IVW results.

To evaluate the robustness of our results, we performed extensive sensitivity analyses. The MR-Egger intercept test assessed potential horizontal pleiotropy (Bowden et al., 2015), while Cochran’s Q test evaluated the heterogeneity among instrumental variables (Bowden et al., 2018). MR pleiotropy residual sum and outlier (MR-PRESSO) analysis was also implemented to identify and exclude outlying instruments based on their contribution to heterogeneity (Verbanck et al., 2018). Any identified outliers were removed and a corrected IVW estimate was recalculated. Funnel plots were generated to visually inspect the heterogeneity in IVW and MR-Egger regressions. Furthermore, leave-one-out analyses were conducted to examine whether individual IVs significantly influenced the overall findings. Instrument strength was assessed using F-statistics, and if IVs exhibited F-statistics below 10, they were excluded to minimise weak instrument bias. We also applied Steiger filtering to retain only variants explaining more variance in the exposure than in the outcome, thereby enforcing the correct causal direction. To limit regression-dilution from NO Measurement Error (NOME) violations, we required high instrument strength for MR-Egger (I² > 0.90) (Bowden et al., 2016). We controlled the FDR at 5% using the Benjamini-Hochberg procedure across 22 comparisons (p_FDR_ < 0.05).

## Results

### UK Biobank Sample

Descriptive statistics for the 163,733 UKB participants with full chronic pain and PHQ-9 phenotype data from the EoP questionnaire data are shown in Table 1. Before analysis, 3,246 participants were excluded for missing chronic pain or depressive symptom data. Overall, 55.0% reported current chronic pain. Compared with chronic pain-free participants, those with chronic pain were older and more often women (both p < 0.001) and had higher PHQ-9 scores (mean 4.0 vs 1.8; p < 0.001). Exact 1:1 age- and sex-matching removed a further 21,055 participants, yielding a final sample of 142,688 individuals: 71,344 chronic pain cases and 71,344 pain-free controls.

**Table 1.** Descriptive statistics of the unmatched Experience of Pain Questionnaire UK Biobank sample.

| Variable | No Chronic Pain<br>N = 71,934 | Chronic Pain<br>N = 91,799 | p-value |
| --- | --- | --- | --- |
| Female, n (%) | 37,433 (52%) | 55,368 (60%) | <0.001 |
| PHQ-9 Score, mean (SD) | 1.8 (2.7) | 4.0 (4.5) | <0.001 |
| Age, mean (SD) | 65.5 (7.7) | 66.0 (7.6) | <0.001 |

P-values are from two-sided tests: Pearson's $\chi^2$ test for the categorical variable (sex) and Wilcoxon rank-sum test for continuous variables (age, PHQ-9).

### Depressive symptom prevalence in chronic pain

Compared to participants without chronic pain, those with chronic pain showed a higher prevalence of depressive symptoms across all severity levels and individual items (all p_FDR_ < 0.001; Figure 1). The severity distribution was shifted upward, with prevalence ratios (PRs) of 2.66 for mild, 4.52 for moderate, 5.71 for moderately severe, and 7.50 for severe symptoms. Cardinal symptoms of depression, anhedonia and depressed mood were 4.00 and 3.64 times more prevalent in chronic pain, respectively. All other symptoms were at least 2.79 times more common in the chronic pain group, with the largest proportional increase in psychomotor changes (PR = 6.60).

**Figure 1.**
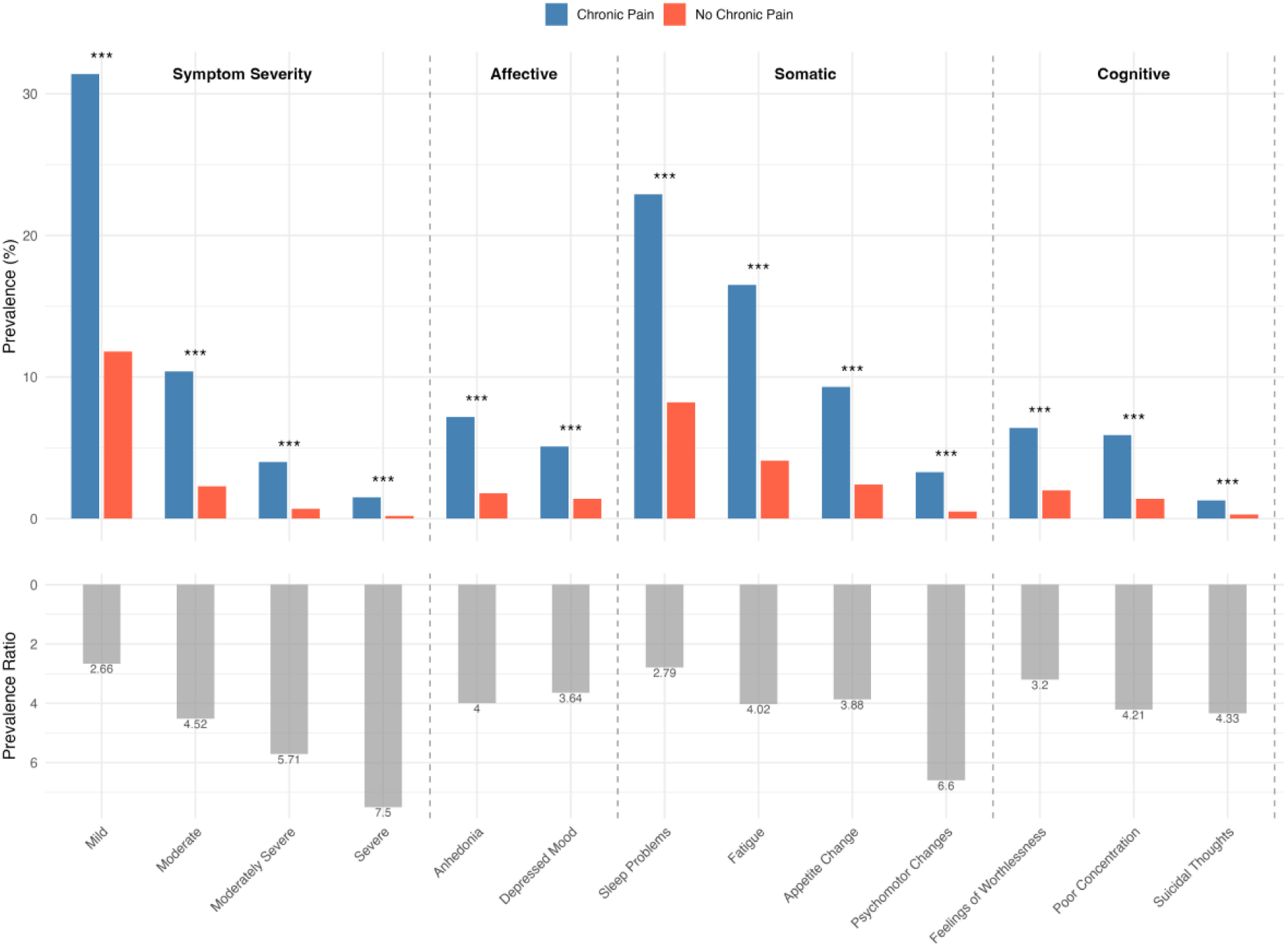
Prevalence of depressive symptom severity and individual symptoms by chronic pain status. Top panel: Prevalence (%) of depression symptom severity and individual depression symptoms in chronic pain cases (blue) and controls (red). Asterisks denote statistical significance after FDR correction for multiple comparisons (*** = p_FDR_ < 0.001). Bottom panel: Prevalence ratios (prevalence in chronic pain/prevalence in no chronic pain) for each item. N = 142,688.

### Genetic correlation of chronic pain and depressive phenotypes

Genetic correlation analyses showed that MCP was positively genetically correlated with MDD and each depression symptom (all p_FDR_ < 0.001; Figure 2). The strongest symptom-level correlations were with insomnia, hypersomnia, and appetite/weight gain (r_g_ = [0.50,0.55]). Moderate correlations were observed for anhedonia (r_g_ = 0.47), concentration problems (r_g_ = 0.42), suicidal thoughts (r_g_ = 0.41), appetite/weight loss (r_g_ = 0.41), depressed mood (r_g_ = 0.38), and fatigue/low energy (r_g_ = 0.36). A weaker but significant correlation was found for feelings of worthlessness or guilt (r_g_ = 0.24).

**Figure 2.**
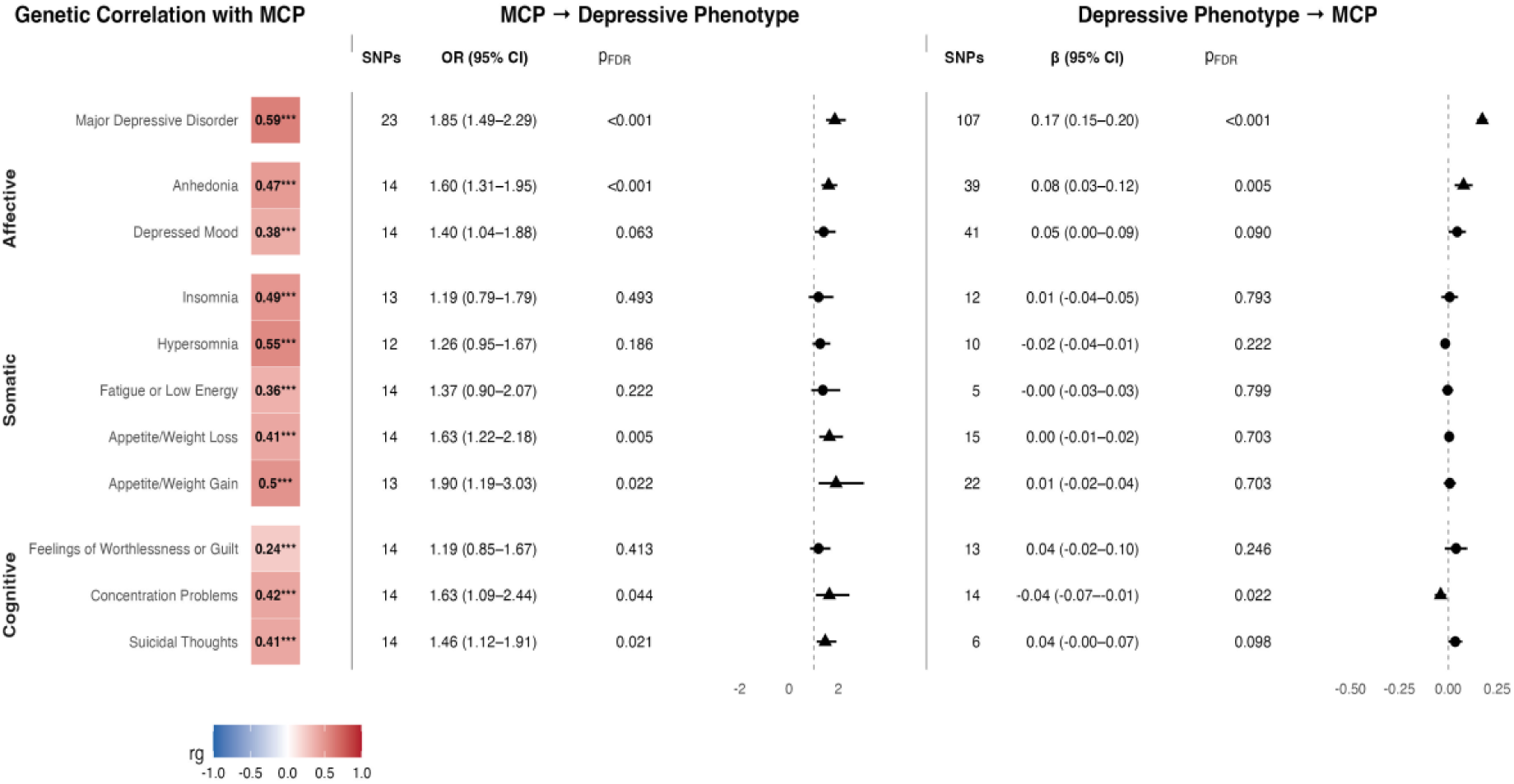
Genetic overlap and directional effects between MCP and depressive phenotypes (MDD and depression symptoms). Left panel shows genetic correlations (rg) between MCP and each depressive phenotype. Asterisks denote statistical significance after FDR correction for multiple comparisons (*** = p_FDR_ < 0.001). Middle panel displays two-sample Mendelian randomisation (MR) estimates for the effect of genetic liability to MCP on each depressive phenotype; right panel shows the reverse direction. Effect sizes are presented as odds ratios (OR) with 95% confidence intervals (CIs) when the outcome was binary (MDD and depressive symptoms), and as β coefficients with 95% CIs when the outcome was continuous (MCP). Triangles indicate significant results (p_FDR_ < 0.05). MDD = major depressive disorder, MCP = multisite chronic pain.

### Causal effects between chronic pain and depressive phenotypes

To identify potential causal associations between chronic pain and depression and symptoms of depression, we conducted a bidirectional MR analysis (Figure 2). Using IVW analysis, we identify significant positive associations between genetic liability to MCP and increased risk of MDD (OR = 1.85, p_FDR_ < 0.001), anhedonia (OR = 1.60, p_FDR_ < 0.001), appetite/weight loss (OR = 1.63, p_FDR_ = 0.006), appetite/weight gain (OR = 1.90, p_FDR_ = 0.022), concentration problems (OR = 1.63, p_FDR_ = 0.044) and suicidal thoughts (OR = 1.46, p_FDR_ = 0.021).

In the reverse direction, we also found genetic liability to MDD (β = 0.17, p_FDR_ < 0.001) and anhedonia (β = 0.08, p_FDR_ = 0.005) to have a significant positive association with MCP. Our MR findings also identified genetic liability to concentration problems to be significantly negatively associated with MCP (β = -0.04, p_FDR_ = 0.022).

Significant IVW estimates were supported by consistent estimates from complementary MR methods (Supplementary Table 6), and additional sensitivity analyses supported the robustness of our findings. Although Cochran’s Q statistic indicated heterogeneity among some of the genetic instruments (Supplementary Table 8), the MR-Egger intercept test showed no evidence of horizontal pleiotropy in any significant MR analyses (Supplementary Table 7). Of the significant IVW estimates, MR-PRESSO analysis revealed five and one outliers in MCP → MDD and anhedonia → MCP analyses, respectively (Supplementary Table 9). However, upon exclusion of outliers and recalculating the IVW analyses, causal estimates were not found to have changed significantly. Additionally, funnel plots for each analysis appeared symmetrical (Supplementary Figures 1–22), and leave-one-out analyses demonstrated that the IVW estimates were not substantially influenced by any single variant (Supplementary Figures 23–44).

## Discussion

Findings from our observational analysis demonstrate a strong association between chronic pain and the overall severity and individual occurrences of depressive symptoms. There was an increased prevalence of all symptom severity levels in chronic pain, however, the prevalence ratio widened with increasing severity, culminating in a 7.5-fold increase of severe depressive symptoms in those with chronic pain. These findings are consistent with other work previously demonstrating the increase in depression severity when both conditions co-occur (Bair et al., 2003). Recognising the marked increase in severity underscores the importance of better understanding the mechanisms linking these disorders to provide more effective treatment.

To our knowledge, item-level PHQ-9 symptom prevalences in chronic pain have not been previously reported in UKB or other large cohorts. In addition, previous association studies in smaller samples have evaluated the prevalence and network dynamics of individual symptoms of depression in only those with chronic pain (McWilliams et al., 2017), or have included pain as a node in depression symptom networks (Yang et al., 2022). By contrast, in this study, by quantifying the prevalence of individual depressive symptoms by chronic pain status, we not only identify which symptoms of depression are the most common in chronic pain but also which show the largest proportional increase given the presence of chronic pain. Therefore, our observational analysis yields key insights: every depressive symptom is more prevalent in chronic pain, with each showing at least a 2.79-fold increase; sleep problems, fatigue, and appetite change are the most commonly endorsed symptoms in both those with and without chronic pain; and psychomotor change, suicidal thoughts, and concentration problems exhibit the largest proportional increases given chronic pain. Overall, these findings reiterate the substantial depressive symptom burden experienced by those with chronic pain and highlight specific symptoms for screening and treatment.

We defined symptom-level cases as those reporting a symptom for at least “more than half the days”, to capture persistent, clinically meaningful symptoms and to align with DSM-5 criteria that several symptoms occur “nearly every day” (American Psychiatric Association, 2013). This conservative cutoff likely increases specificity, possibly reducing false positives from transient distress or pain-related somatic overlap, but also reduces sensitivity and may lower absolute prevalence estimates by classifying “several days” responses as controls. Whether any misclassification differs between those with and without chronic pain is uncertain; evaluating alternative cut-points in future work would clarify threshold effects.

When assessing the prevalence of depressive symptoms in chronic pain, it is critical to consider the source of these associations. A study investigating how patients with chronic pain interpreted and responded to the PHQ-9 found that several problematic items limited the construct validity of the questionnaire, increasing the risk of inflated depression scores (Aagaard et al., 2023). An example from our results is the increased prevalence of psychomotor changes, which had the largest prevalence ratio of all symptoms in our results, which may be partially explained by decreased mobility and heightened restlessness in the context of chronic pain (Hoogwout et al., 2015; Ogawa et al., 2020). Therefore, it is likely that the symptom overlap between depression and chronic pain (and other related comorbidities) partially explains the strong phenotypic associations observed in this study.

In addition to symptom overlap, there is likely to be a complex network of variables promoting the associations we have observed between chronic pain and individual depressive symptoms. Through sample matching, our analysis controlled for the effects of sex and age, important risk factors for both chronic pain (Dagnino & Campos, 2022) and depression (Zenebe et al., 2021), however other potential confounders, such as biological, socioeconomic, or lifestyle factors, were not accounted for, meaning the observed associations may not be independent of these influences. These considerations, together with the symptom overlap noted above, underscore the value of our genetic approach. By leveraging germline genetic variants, which are fixed at conception, our genetic analyses mitigate recall bias. Furthermore, MR analysis allows us to estimate causal effects between chronic pain and individual symptoms of depression, helping distinguish association from putative causation.

Findings from our genetic correlation analysis demonstrated a substantial positive correlation between MCP and MDD, comparable to that shown by Johnston et al using an earlier MDD GWAS (r_g_ = 0.53) (Johnston et al., 2019). Depression symptom-level genetic correlations with MCP were all positive and significant and ranged from low (feelings of worthlessness: r_g_ = 0.25) to moderate (hypersomnia: r_g_ = 0.55). The significant positive correlations observed suggest a shared genetic architecture and help explain the associations identified in our observational analysis. This genetic overlap may reflect horizontal pleiotropy, in which specific genetic variants independently increase the risk for both conditions. Alternatively, it may indicate vertical pleiotropy, which we have explored in our MR analysis, where the genetic influence of one trait contributes causally or directionally to the development of the other (Pingault et al., 2018).

In addition to supporting a bidirectional causal relationship between MCP and MDD, our symptom-level MR analyses also indicated a bidirectional causal association between MCP and anhedonia. Within the framework of the network theory of psychopathology, which defines depression as a product of its interacting symptoms, rather than a latent factor preceding symptoms (Borsboom, 2017), our MR findings of a bidirectional association between chronic pain and anhedonia suggest that this symptom-level causal pathway may partially drive the broader association observed between chronic pain and depression. This finding highlights anhedonia as an important treatment target for patients with co-occurring chronic pain and depression.

We also found evidence consistent with a causal pathway from MCP to additional symptoms: appetite/weight loss, appetite/weight gain, concentration problems, and suicidal thoughts. Clarifying the direction of these effects helps explain the phenotypic correlations in our observational analyses and may indicate a symptom-network mechanism whereby chronic pain triggers specific depressive symptoms that, through cascading interactions (Schlechter et al., 2023), culminate in depression. Finally, we observed an unexpected MR estimate consistent with concentration problems exerting a protective effect on MCP. Although cognitive difficulties (including concentration problems) are frequently among the most burdensome features in chronic pain populations (Geisser & Kratz, 2018), one possible mechanism is that reduced ability to sustain attention may limit ruminative focus on persistent pain, which is known to exacerbate pain duration and severity (Fonseca das Neves et al., 2023).

One of the major strengths of this study is the use of a large UKB sample in our observational analysis, alongside the current largest available GWAS summary statistics for MDD, MCP, and individual depressive symptoms for each genetic approach. Additionally, disaggregating depression into its constituent symptoms provided more homogenous measures than depression and highlighted specific symptoms as potential targets for screening and treatment. Finally, our methods robustly characterised depressive symptoms in chronic pain from different perspectives, in particular, our genetic analysis mitigated key limitations of observational data (e.g., reverse causality, recall bias), providing complementary explanations for the association observed.

Several limitations should also be addressed. As highlighted previously, depression symptom prevalence is likely inflated when using self-report questionnaires in chronic pain due to symptom overlap between the disorders, limiting what conclusions we can draw from our analyses. Additionally, observational data did not capture neurovegetative symptom polarity (e.g., insomnia vs. hypersomnia), likely removing important insights from our findings. A standard limitation of MR analysis is that it relies on three core assumptions that can be violated by horizontal pleiotropy, weak instruments, and measurement error, biasing estimates (de Leeuw et al., 2022). We used robust estimators and extensive sensitivity analyses to mitigate these risks, but residual bias cannot be ruled out. Finally, our observational analyses were conducted in UKB, a volunteer cohort that is not representative of the UK population, limiting generalisability (Fry et al., 2017; Keyes & Westreich, 2019). Additionally, the GWAS summary statistics also used data collected from non-representative cohorts, the majority including UKB, and were limited to European ancestry, further restricting applicability across ancestries and demographic groups.

We identified overlaps in genetic architecture between chronic pain and depression and its symptoms on a global genomic scale; however, extending this work to identify specific genomic loci shared between both conditions and the implicated genes would provide greater insight into the overlapping biological processes promoting this comorbidity, and may highlight potential novel treatment targets (Li et al., 2025).

## Conclusion

This study found all individual symptoms of depression, across affective, somatic and cognitive domains, to be at least 2.79 times more prevalent in chronic pain. Results from genetic analyses indicate that these associations can be explained, at least in part, by shared genetic architecture between traits and putative causal associations between MCP and several depressive symptoms. Specifically, our findings support a bidirectional causal association between MCP and anhedonia, and a one-way association from MCP liability to several other depressive symptoms. The marked increase in all depressive symptoms among those with chronic pain emphasises the severe and comprehensive depressive symptom burden in those with chronic pain, demonstrating the importance of robust tools for depression screening in chronic pain. Our genetic findings help to explain these observational correlations and highlight specific symptoms that may drive the association between chronic pain and depression.

## Data availability

Data from UK Biobank (application number 4844) were used for the observational association analysis in this study. Researchers can apply to use UK Biobank data for health-related research that is in the public interest. Further information is available from the <u>UK Biobank website</u>. GWAS summary statistics for MDD and individual depressive symptoms can be downloaded from https://pgc.unc.edu/for-researchers/download-results/, and MCP summary statistics are available at https://researchdata.gla.ac.uk/822/.

## Code availability

The custom code used to perform all analyses for this study will be made publicly available after publication at https://github.com/hannahcasey/UKB_chronic_pain_depressive_symptoms.

## Ethics and consent

The UK Biobank study was approved by the North West Multi-centre Research Ethics Committee (REC reference: 11/NW/0382). Written informed consent was obtained from each participant.

## Conflicts of interest

The authors declare no competing interests.

## Grant information

Access to UK Biobank was made possible through the Wellcome Trust Strategic Award “Stratifying Resilience and Depression Longitudinally” (STRADL) (reference 104036/Z/14/Z). HC is funded by the Medical Research Council and the University of Edinburgh through the Precision Medicine Doctoral Training Program.

## Author contributions

**Hannah Casey:** Conceptualisation; data curation; formal analysis; methodology; software; validation; visualisation; writing - original draft; writing - review & editing

**Mark J. Adams:** Data curation, resources

**Andrew M. McIntosh:** Funding acquisition

**Marie T. Fallon:** Conceptualisation; supervision; writing - review & editing

**Daniel J. Smith:** Conceptualisation; supervision; writing - review & editing

**Rona J. Strawbridge:** Conceptualisation; methodology; supervision; writing - review & editing

**Heather C. Whalley:** Conceptualisation; methodology; supervision; writing - review & editing

## Notes

### Competing Interest Statement

The authors have declared no competing interest.

