## Supplementary Figures for "Symptoms of depression in chronic pain: prevalence in UK Biobank and shared genetic factors"

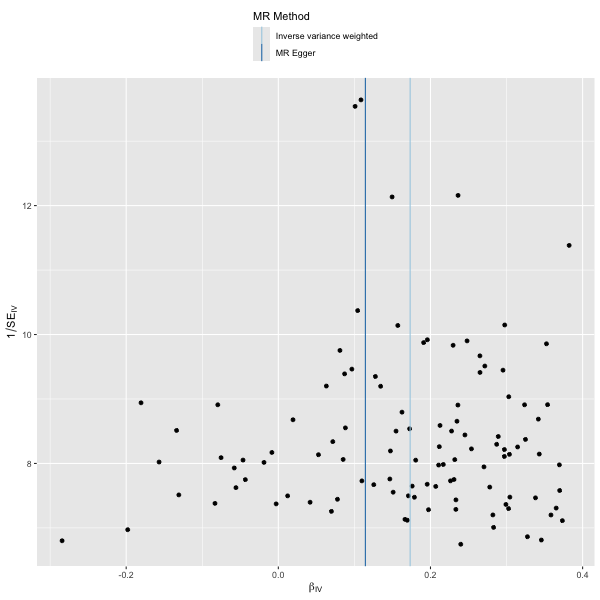

**Supplemental Figure 1.** Funnel plot of MDD on MCP.

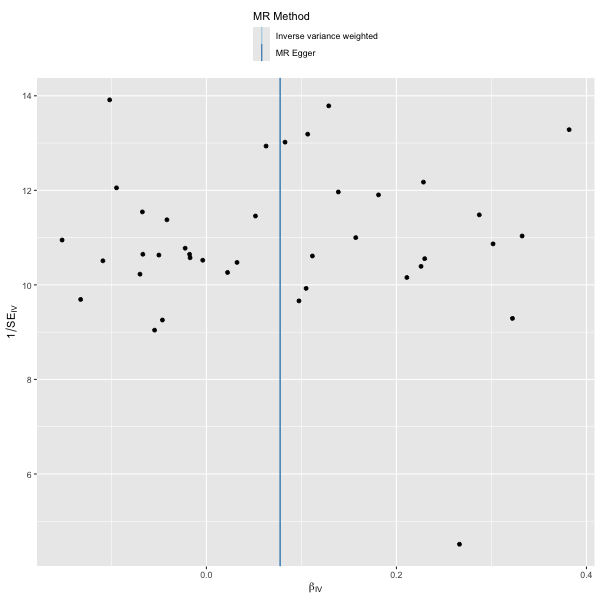

**Supplemental Figure 2.** Funnel plot of anhedonia on MCP.

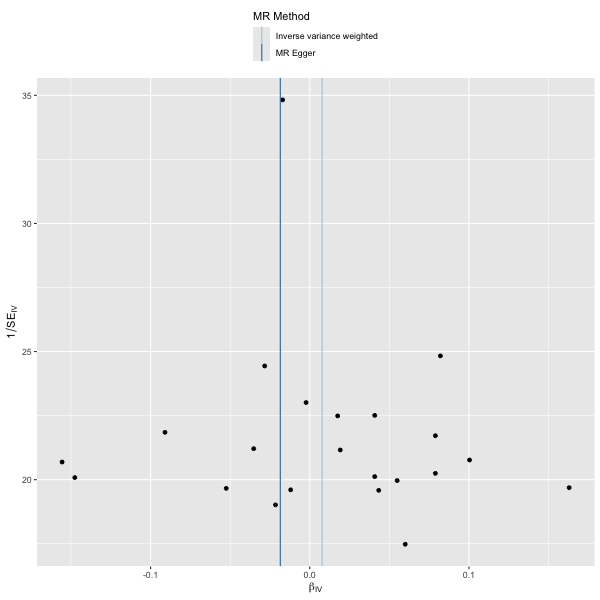

**Supplemental Figure 3.** Funnel plot of appetite on weight gain MCP.

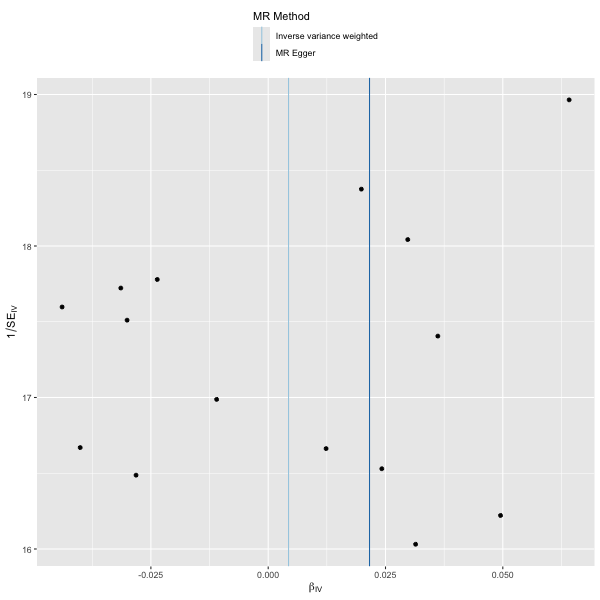

**Supplemental Figure 4.** Funnel plot of appetite on weight loss MCP.

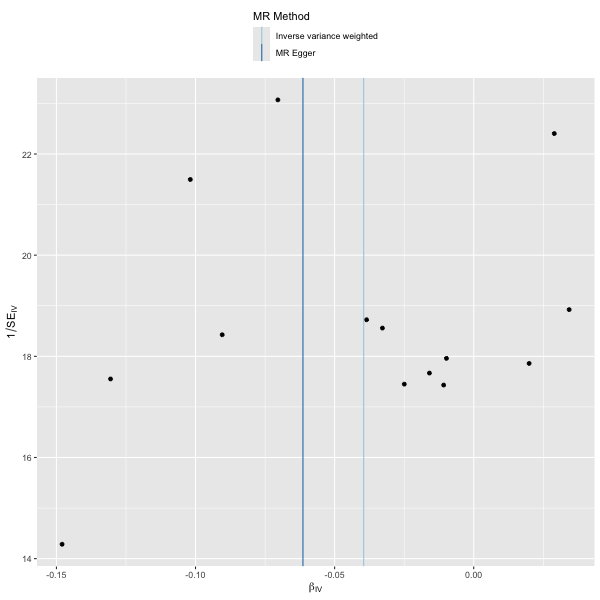

**Supplemental Figure 5.** Funnel plot of concentration on difficulty MCP.

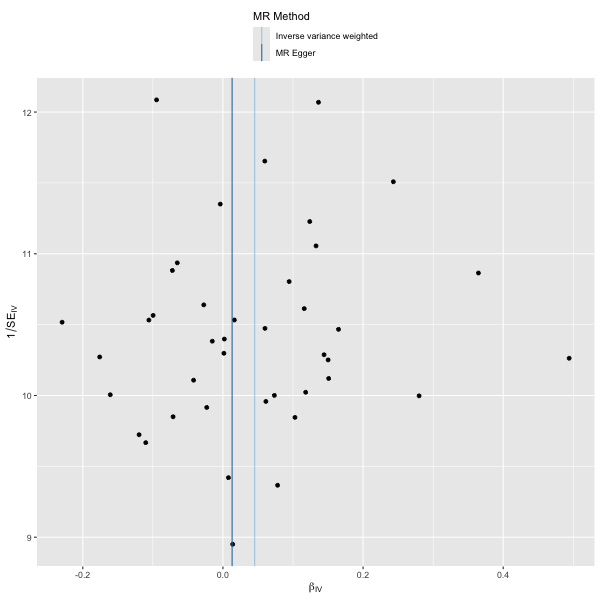

**Supplemental Figure 6.** Funnel plot of depressed on mood MCP.

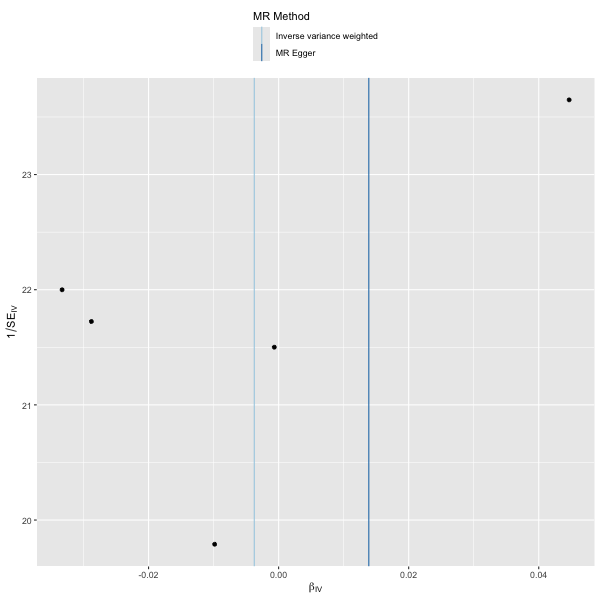

**Supplemental Figure 7.** Funnel plot of fatigue on MCP.

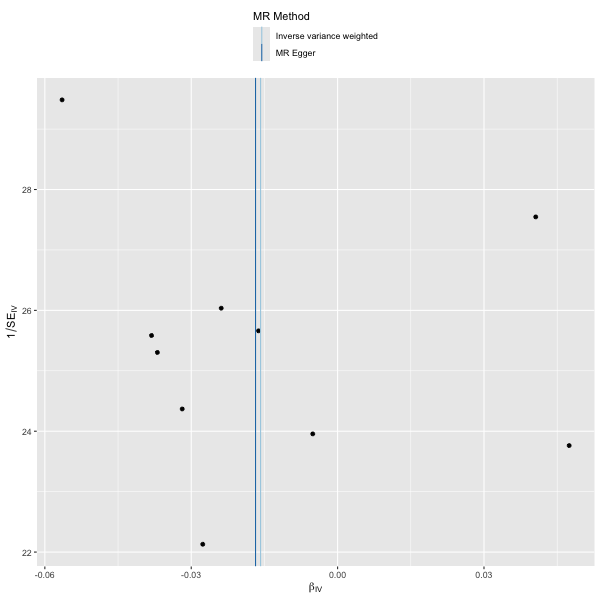

**Supplemental Figure 8.** Funnel plot of hypersomnia on MCP.

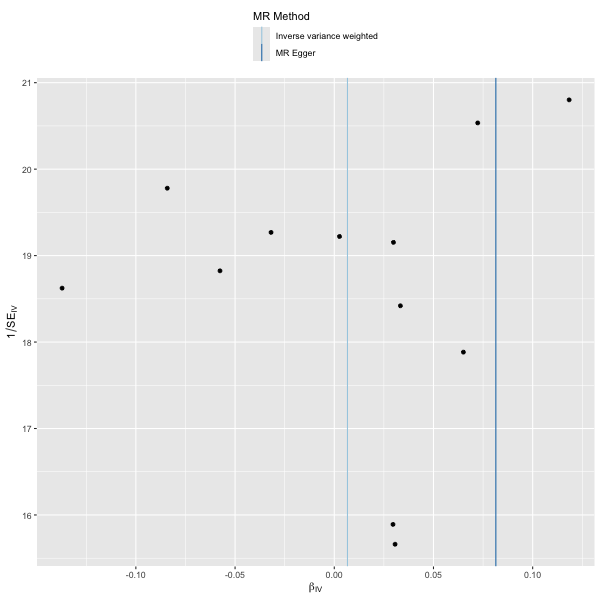

**Supplemental Figure 9.** Funnel plot of insomnia on MCP.

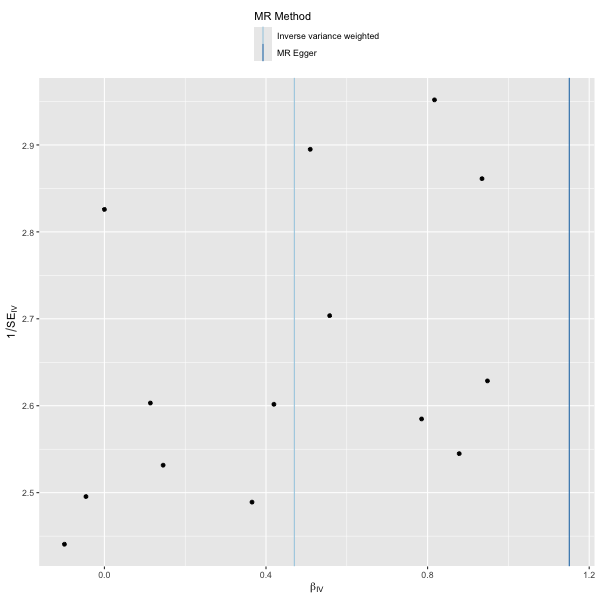

**Supplemental Figure 10.** Funnel plot of MCP on anhedonia.

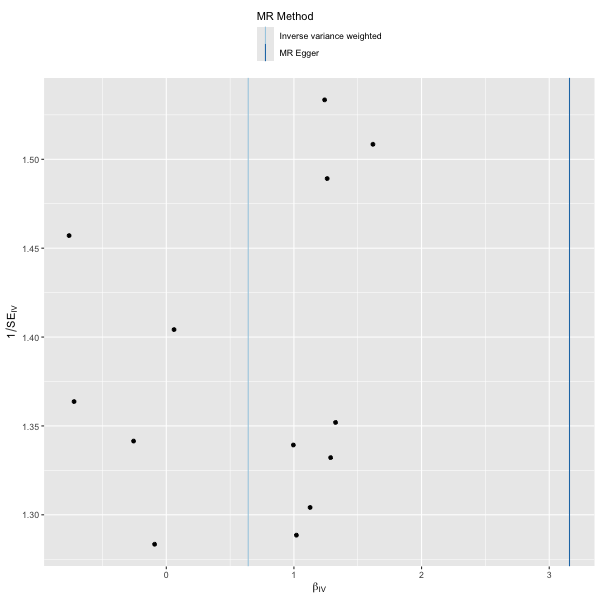

**Supplemental Figure 11.** Funnel plot of MCP on appetite weight gain.

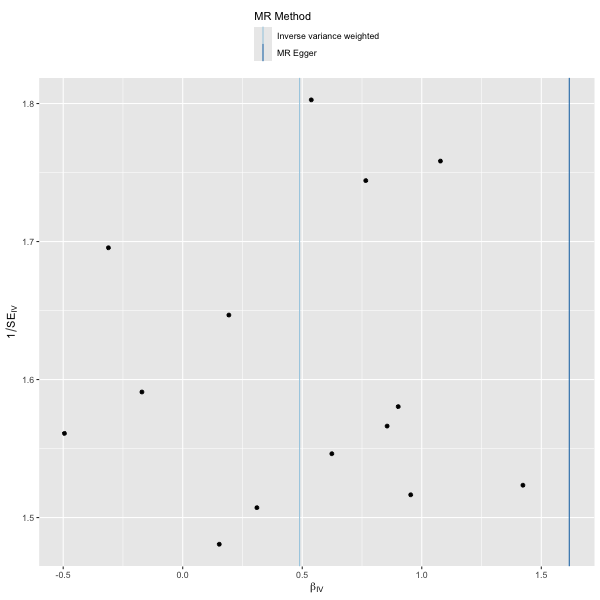

**Supplemental Figure 12.** Funnel plot of MCP on appetite weight loss.

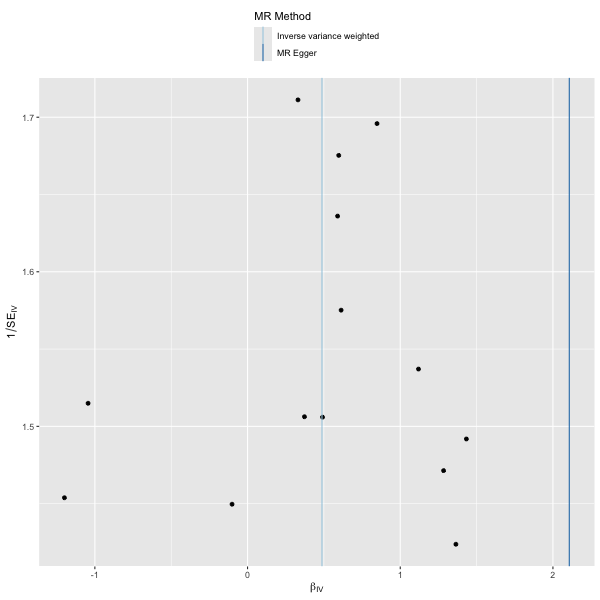

**Supplemental Figure 13.** Funnel plot of MCP on concentration difficulty.

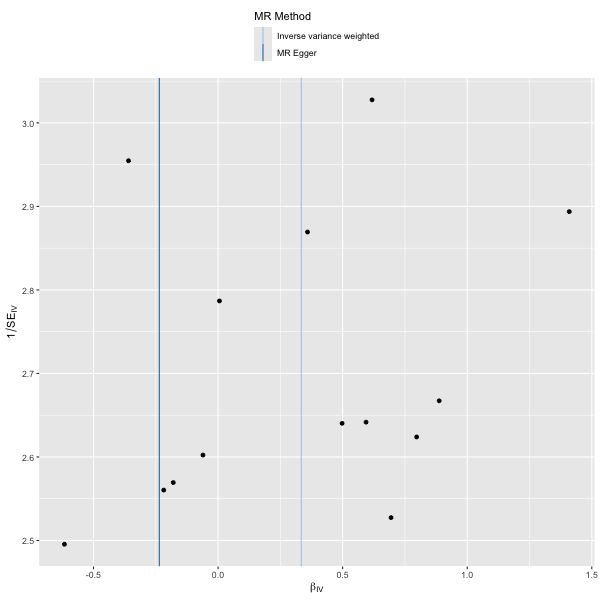

**Supplemental Figure 14.** Funnel plot of MCP on depressed mood.

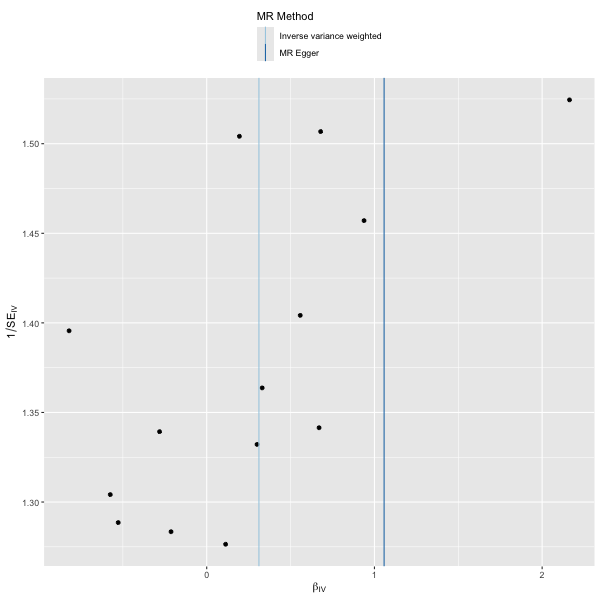

**Supplemental Figure 15.** Funnel plot of MCP on fatigue.

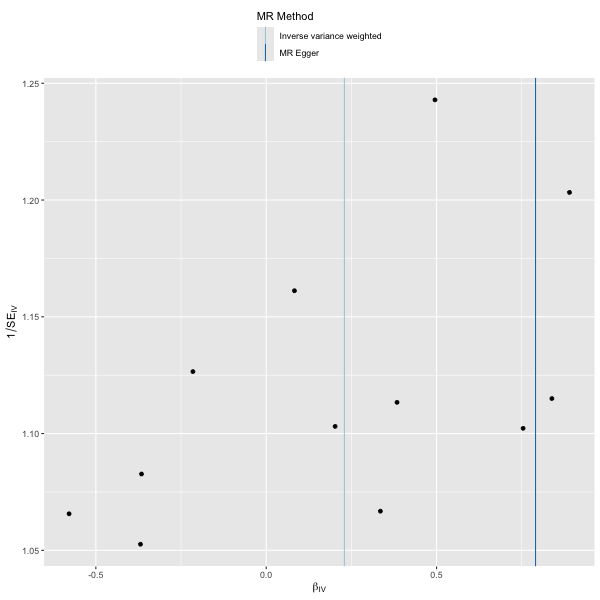

**Supplemental Figure 16.** Funnel plot of MCP on hypersomnia.

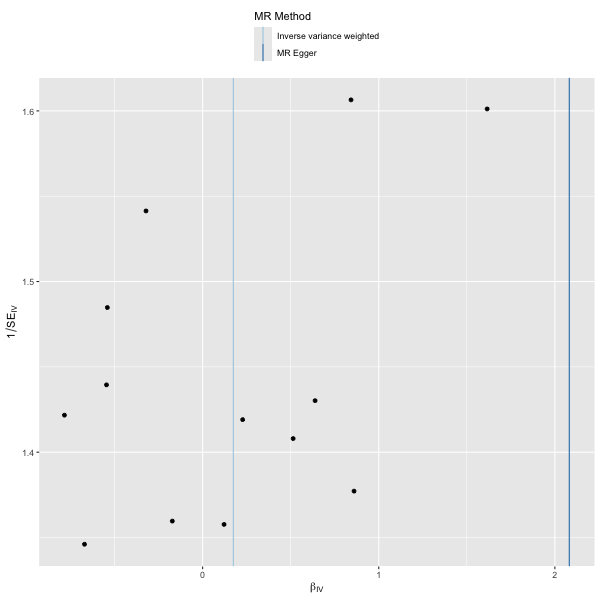

**Supplemental Figure 17.** Funnel plot of MCP on insomnia.

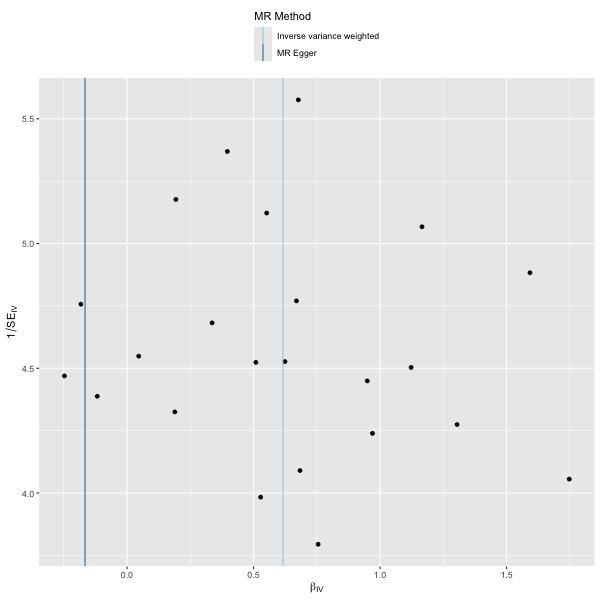

**Supplemental Figure 18.** Funnel plot of MCP on MDD.

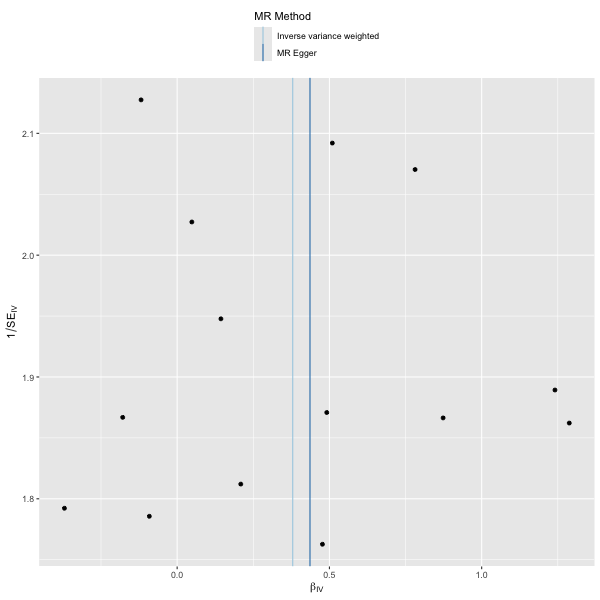

**Supplemental Figure 19.** Funnel plot of MCP on suicidal.

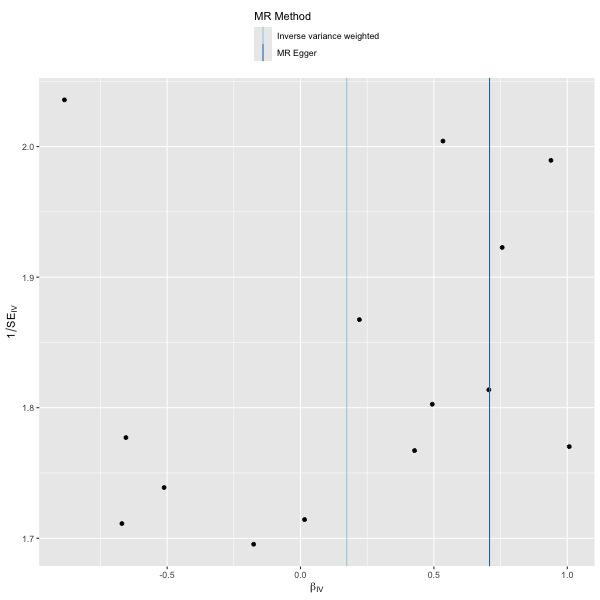

**Supplemental Figure 20.** Funnel plot of MCP on worthless.

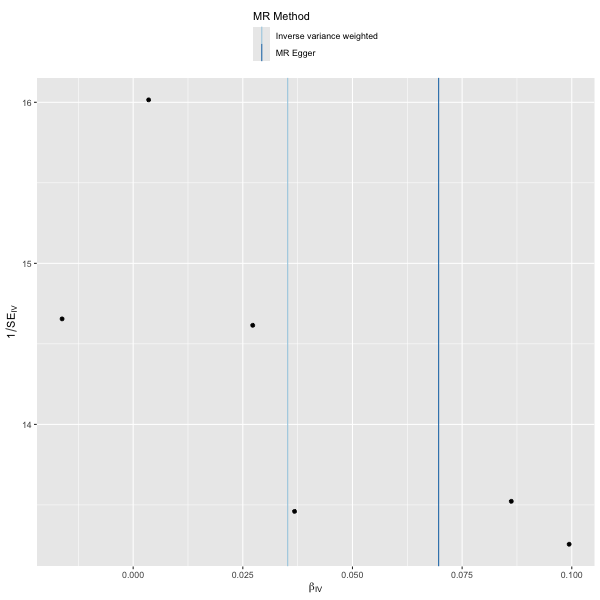

**Supplemental Figure 21.** Funnel plot of suicidal on MCP.

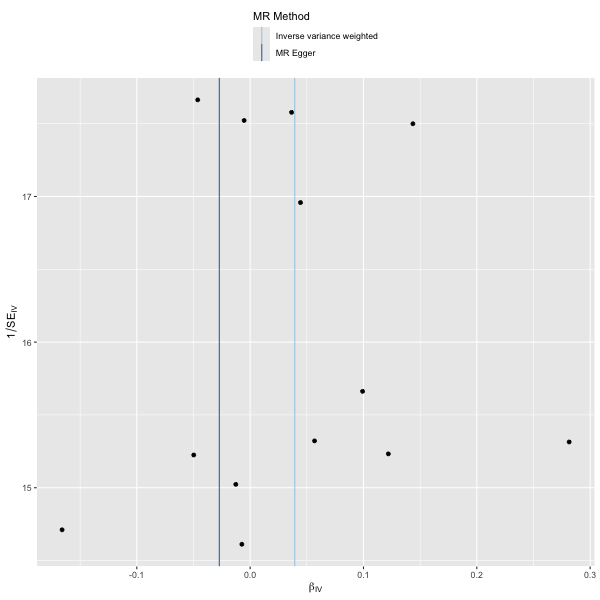

**Supplemental Figure 22.** Funnel plot of worthless on MCP.

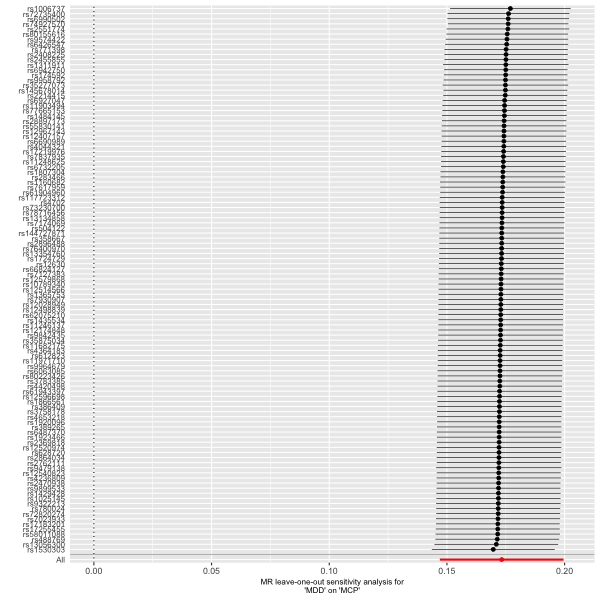

**Supplemental Figure 23.** Leave-one-out plot of MDD on MCP.

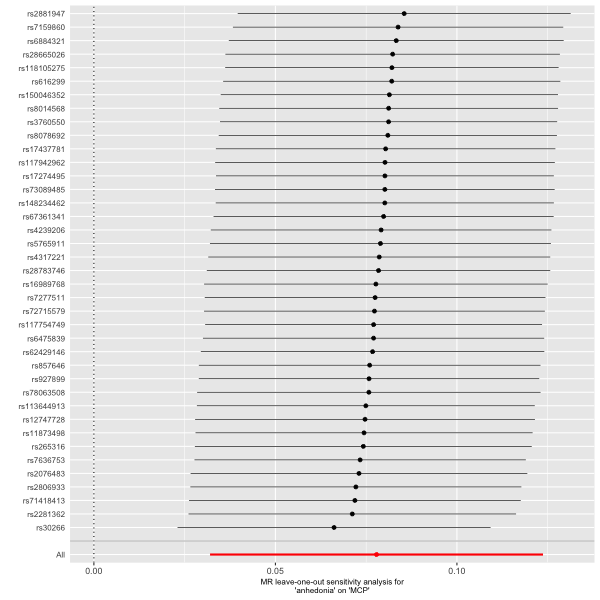

**Supplemental Figure 24.** Leave-one-out plot of anhedonia on MCP.

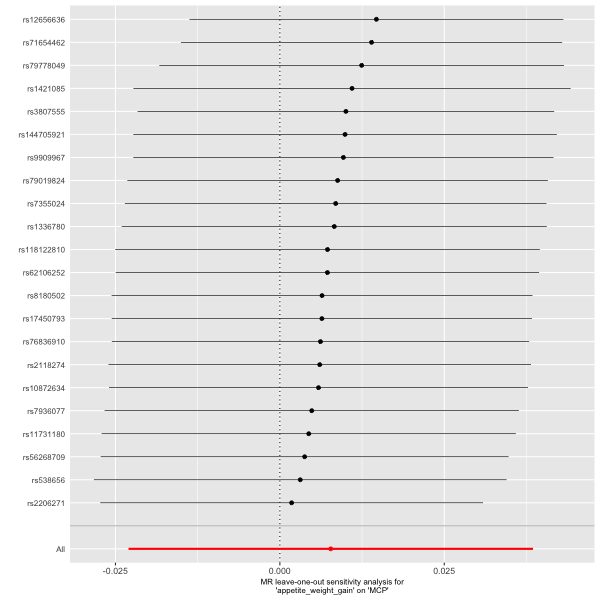

**Supplemental Figure 25.** Leave-one-out plot of appetite on weight gain MCP.

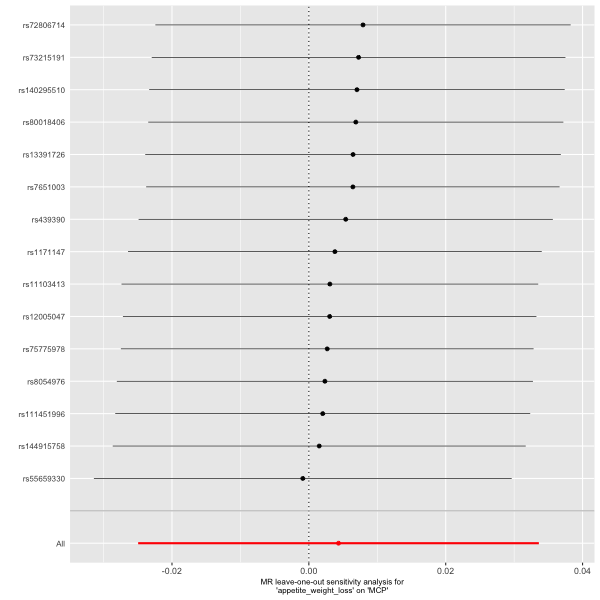

**Supplemental Figure 26.** Leave-one-out plot of appetite on weight loss MCP.

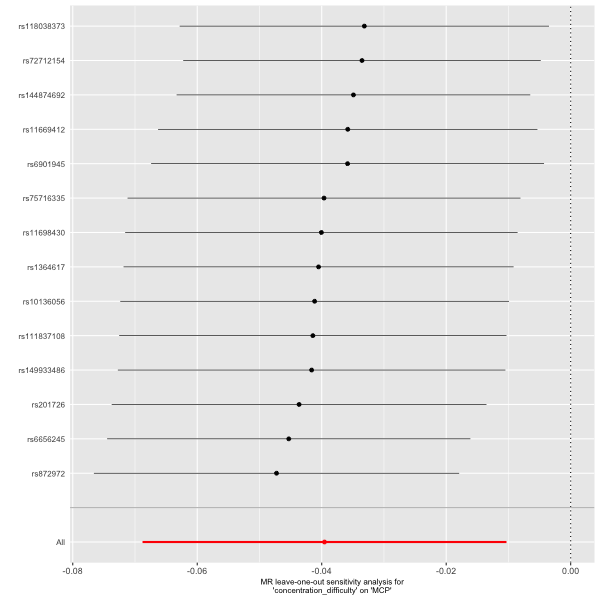

**Supplemental Figure 27.** Leave-one-out plot of concentration on difficulty MCP.

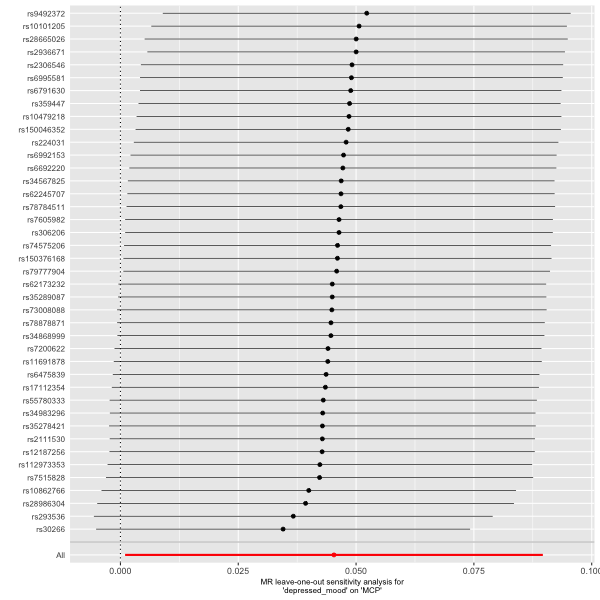

**Supplemental Figure 28.** Leave-one-out plot of depressed on mood MCP.

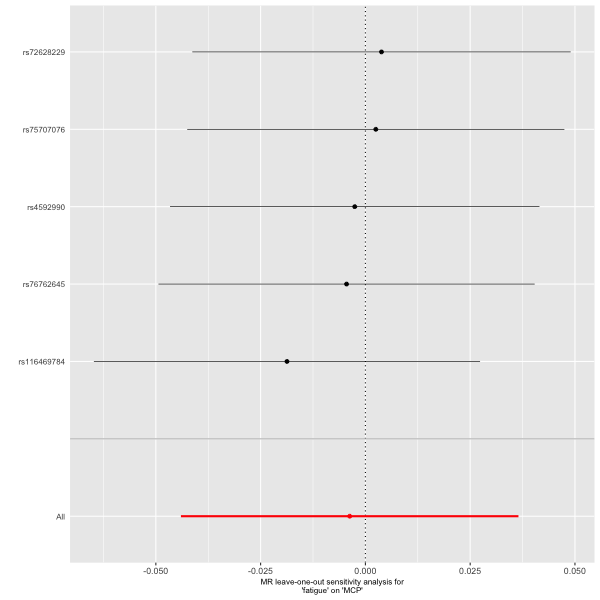

**Supplemental Figure 29.** Leave-one-out plot of fatigue on MCP.

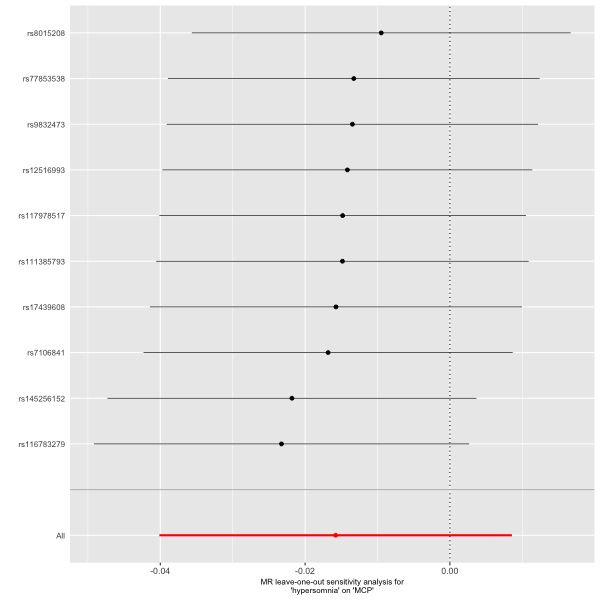

**Supplemental Figure 30.** Leave-one-out plot of hypersomnia on MCP.

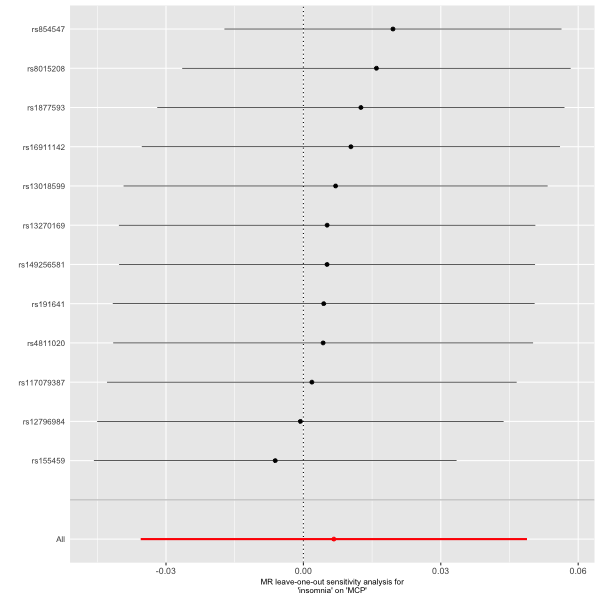

**Supplemental Figure 31.** Leave-one-out plot of insomnia on MCP.

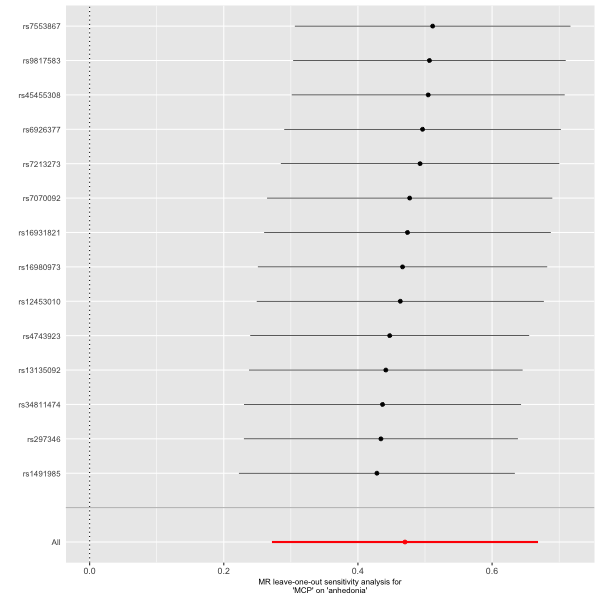

**Supplemental Figure 32.** Leave-one-out plot of MCP on anhedonia.

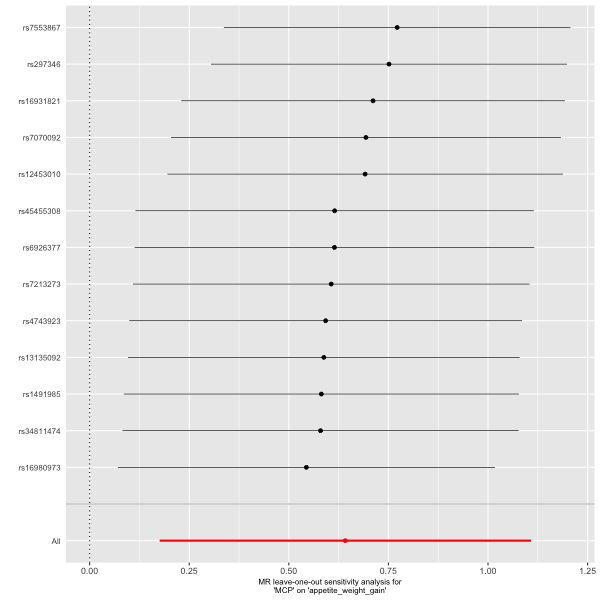

**Supplemental Figure 33.** Leave-one-out plot of MCP on appetite weight gain.

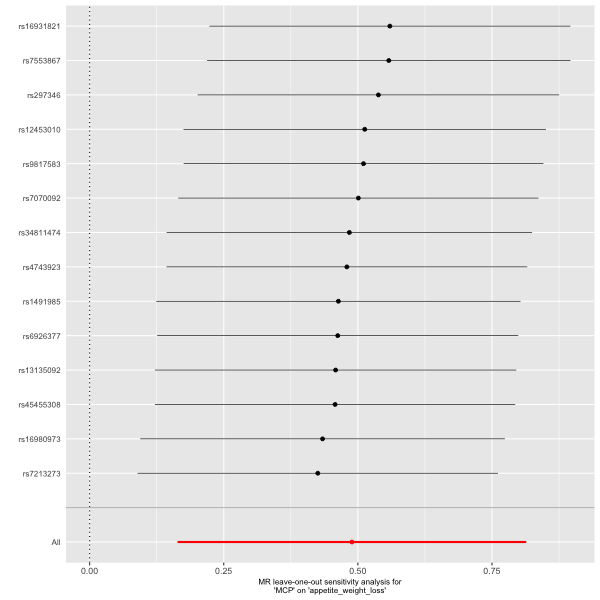

**Supplemental Figure 34.** Leave-one-out plot of MCP on appetite weight loss.

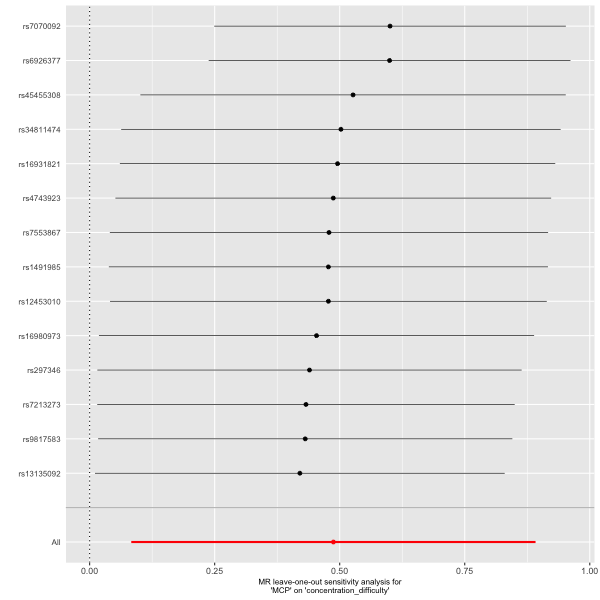

**Supplemental Figure 35.** Leave-one-out plot of MCP on concentration difficulty.

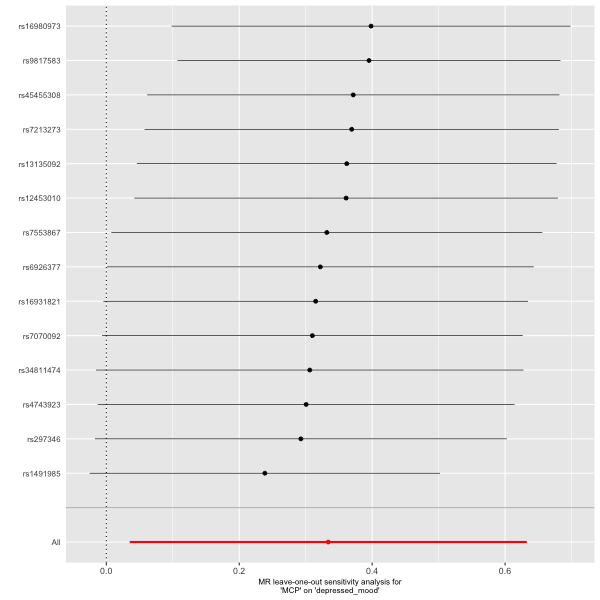

**Supplemental Figure 36.** Leave-one-out plot of MCP on depressed mood.

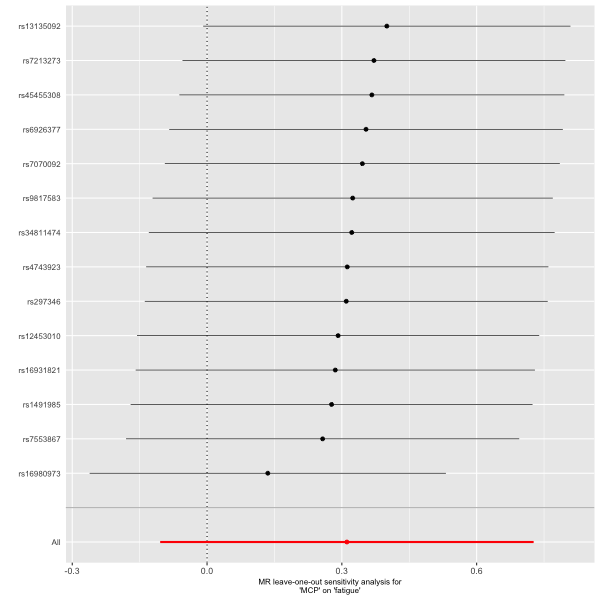

**Supplemental Figure 37.** Leave-one-out plot of MCP on fatigue.

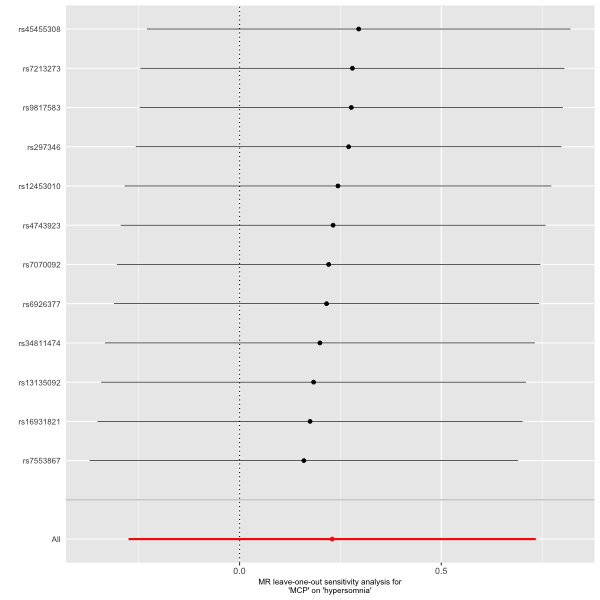

**Supplemental Figure 38.** Leave-one-out plot of MCP on hypersomnia.

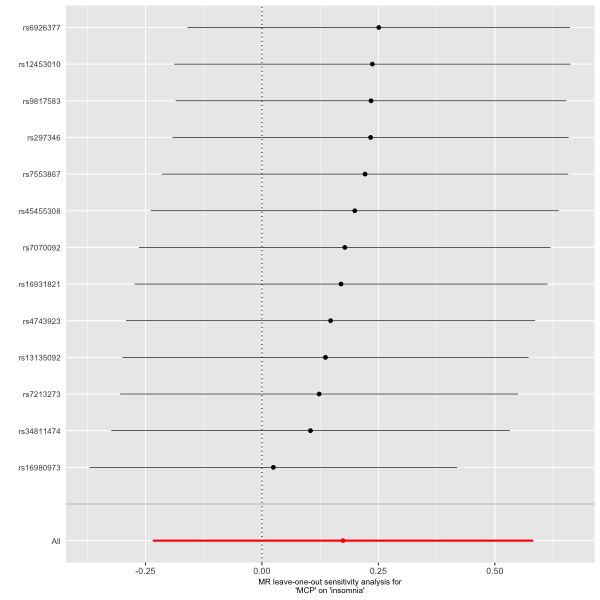

**Supplemental Figure 39.** Leave-one-out plot of MCP on insomnia.

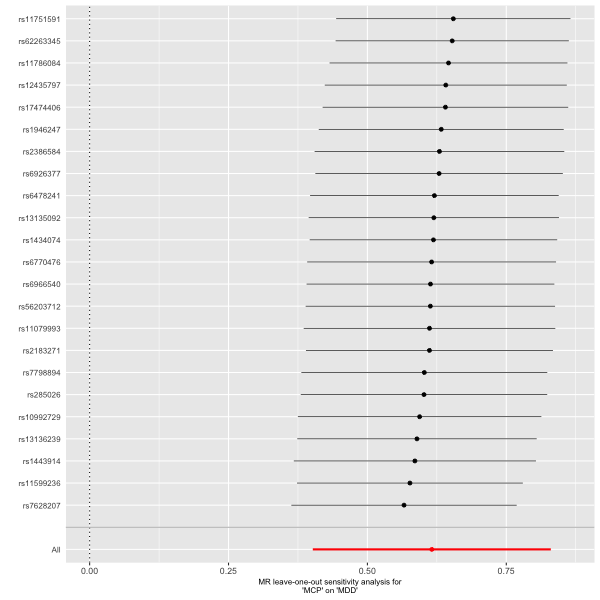

**Supplemental Figure 40.** Leave-one-out plot of MCP on MDD.

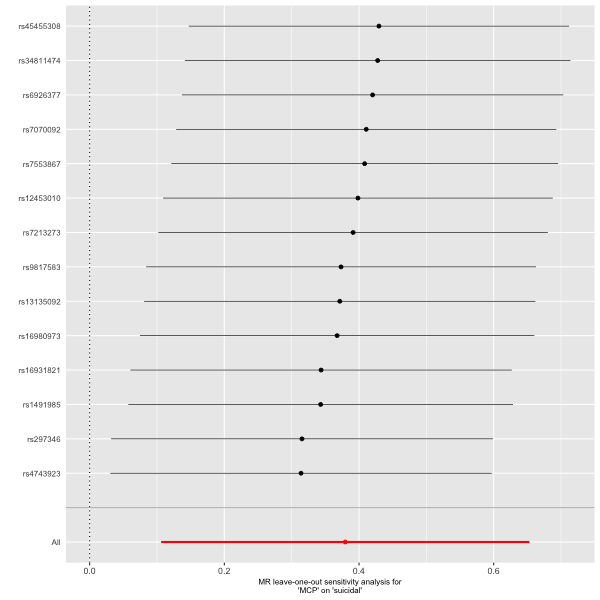

**Supplemental Figure 41.** Leave-one-out plot of MCP on suicidal.

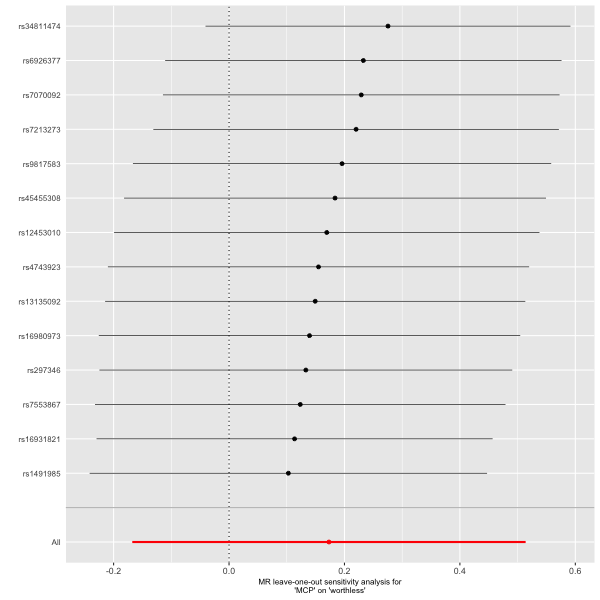

**Supplemental Figure 42.** Leave-one-out plot of MCP on worthless.

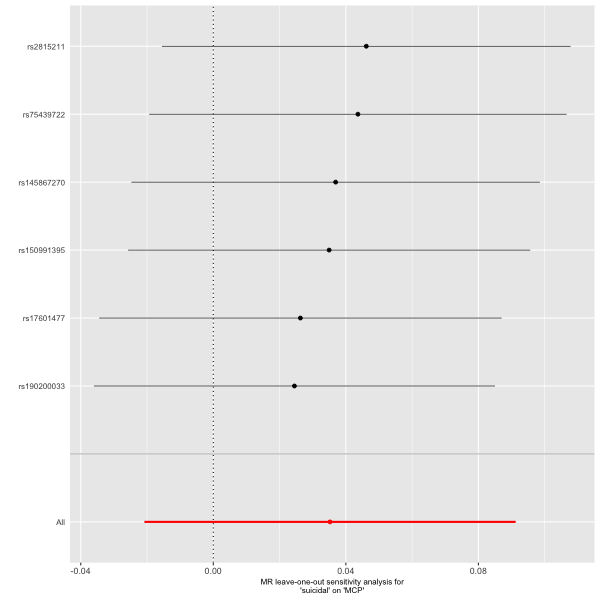

**Supplemental Figure 43.** Leave-one-out plot of suicidal on MCP.

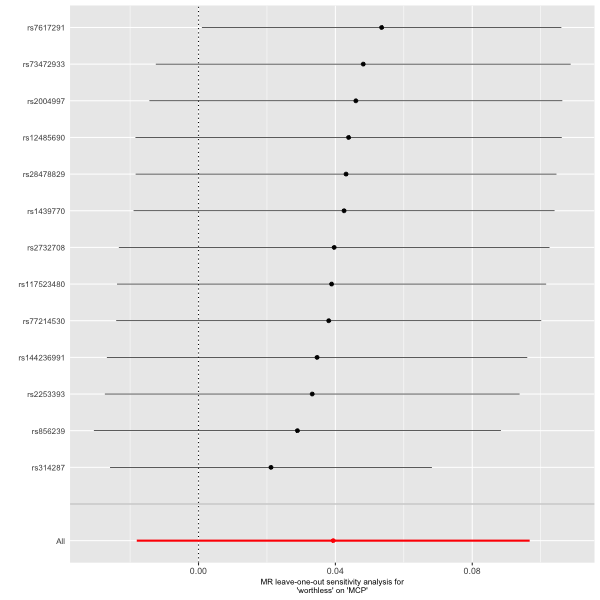

**Supplemental Figure 44.** Leave-one-out plot of worthless on MCP.
